# COVID-19 Survivors’ Reports of the Timing, Duration, and Health Impacts of Post-Acute Sequelae of SARS-CoV-2 (PASC) Infection

**DOI:** 10.1101/2021.03.22.21254026

**Authors:** Natalie Lambert, Survivor Corps, Sarah A. El-Azab, Nathan S. Ramrakhiani, Anthony Barisano, Lu Yu, Kaitlyn Taylor, Alvaro Esperanca, Charles A. Downs, Heather L. Abrahim, Amir M. Rahmani, Jessica L. Borelli, Rana Chakraborty, Melissa D. Pinto

## Abstract

**IMPORTANCE:** Post-Acute Sequelae of SARS-CoV-2 Infection (PASC) is a major public health concern. Studies suggest that 1 in 3 infected with SARS-CoV-2 may develop PASC, including those without initial symptoms or with mild COVID-19 disease.^1, 2^

**OBJECTIVE:** To evaluate the timing, duration, and health impacts of PASC reported by a large group of primarily non-hospitalized COVID-19 survivors.

**DESIGN, SETTING, AND PARTICIPANTS:** A survey of 5,163 COVID-19 survivors reporting symptoms for more than 21 days following SARS-CoV-2 infection. Participants were recruited from Survivor Corps and other online COVID-19 survivor support groups.

**MAIN OUTCOMES AND MEASURES:** Participants reported demographic information, as well as the timing, duration, health impacts, and other attributes of PASC. The temporal distribution of symptoms, including average time of onset and duration of symptoms were determined, as well as the perceived distress and impact on ability to work.

**RESULTS:** On average, participants reported 21.4 symptoms and the number of symptoms ranged from 1 to 93. The most common symptoms were fatigue (79.0%), headache (55.3%), shortness of breath (55.3%), difficulty concentrating (53.6%), cough (49.0%), changed sense of taste (44.9%), diarrhea (43.9%), and muscle or body aches (43.5%). The timing of symptom onset varied and was best described as happening in waves. The longest lasting symptoms on average for all participants (in days) were “frequently changing” symptoms (112.0), inability to exercise (106.5), fatigue (101.7), difficulty concentrating (101.1), memory problems (100.8), sadness (99.2), hormone imbalance (99.1), and shortness of breath (96.9). The symptoms that affected ability to work included the relapsing/remitting nature of illness (described by survivors as “changing symptoms”), inability to concentrate, fatigue, and memory problems, among others. Symptoms causing the greatest level of distress (on scale of 1 “none” to 5 “a great deal”) were extreme pressure at the base of the head (4.4), syncope (4.3), sharp or sudden chest pain (4.2), brain pressure (4.2), headache (4.2), persistent chest pain or pressure (4.1), and bone pain in extremities (4.1).

**CONCLUSIONS AND RELEVANCE:** PASC is an emerging public health priority characterized by a wide range of changing symptoms, which hinder survivors’ ability to work. PASC has not been fully characterized and the trajectory of symptoms and long-term outcomes are unknown. There is no treatment for PASC, and survivors report distress in addition to a host of ongoing symptoms. Capturing patient reports of symptoms through open-ended inquiry is a critical first step in accurately and comprehensively characterizing PASC to ensure that medical treatments and management strategies best meet the needs of individual patients and help mitigate health impacts of this new disease.

## Introduction

The long-term effects of infection with SARS-CoV-2 are poorly understood. This is particularly alarming given that over 118,000,000 persons have contracted SARS-CoV-2 since the virus emerged in late 2019.^3^ Recent studies suggest that nearly a third of individuals having had SARS-CoV-2 infection, including those who are initially asymptomatic, go on to develop symptoms that appear to last for a yet undefined period of time.^1^ These COVID-19 survivors report a wide range of long-lasting sequelae,^4^ some of which are ongoing nine months after infection.^5^

To date most of the research involving SARS-CoV-2 and its consequences has involved the 10% of individuals with severe illness that required hospitalization,^6–8^ leaving a significant gap in our understanding of the health impacts of SARS-CoV-2 infection among the majority (90%) of persons who have been infected to date – those that did not require hospitalization. As data emerged, persons with persistent symptoms 21 or more days following infection have been labeled as “long-haulers,” or those with Post-Acute Sequelae of SARS-CoV-2 (PASC). Not only is public awareness surrounding this topic low, but research concerning PASC is minimal.^9^ Here, we characterize PASC among a participant group composed primarily of non-hospitalized COVID-19 survivors (n=5,163) through a survey that captures symptom timing, duration, and health impacts of PASC.

## Methods

Data for this project were procured through an IRB-approved study and was made possible by a collaboration between the Indiana University School of Medicine and Survivor Corps. Survivor Corps is a COVID-19 grassroots organization and Facebook group dedicated to supporting COVID-19 survivors and scientific research of COVID-19. In August 2020, a REDCap survey was disseminated to Survivor Corps group members on Facebook and in other online COVID-19 groups. The survey was sent out again in November 2020 to collect more data. The purpose of the survey was to understand respondents’ experiences with the timing and duration of the COVID-19 symptoms and how they were impacted by these symptoms.

To participate in the study, all respondents had to be 18 years or older and have been diagnosed with COVID-19 through a positive SARS-CoV-2 antigen or RT PCR test, physician diagnosis, self-diagnosis, or some other method (e.g., positive antibody test). The survey collected self-reported demographic information, an extensive medical history (including underlying conditions), and experiences with COVID-19 (including hospitalization). In addition, the survey listed and asked questions about 101 distinct COVID-19 symptoms and gave respondents the option to report additional symptoms they had experienced that were not listed. The initial survey collected 5,875 responses.

All survey data were manually reviewed and cleaned for analysis. While asymptomatic respondents could participate, only data from respondents who reported symptoms were included in the results. To focus on the experiences of people with PASC, the data analyzed were further limited to respondents who had experienced symptoms for longer than 21 days. The final dataset for analysis consisted of 5,163 participants.

For each symptom, the percentage of participants reporting the symptom, average pain or discomfort associated with the symptom, average duration, and average time to symptom onset were calculated. Means were calculated for participants’ responses to a 5-point Likert scale from “Not at all” to “Very much” for questions about the pain and discomfort, work impairment, and social relationship impact caused by each symptom. Additionally, the percentage of participants that reported the symptom as ongoing and the percentage who reported the symptom as intermittent was calculated for each symptom. The ggplot R package was used to visualize the relationship between symptom discomfort and average duration, the distribution of the number of symptoms reported by each participant, and to create a table containing all symptom metrics calculated for this study.

## Results

### Survey demographics

A total of 5,875 people responded to the survey. Of these, 5,163 reported persistent symptoms. Participants were predominantly White (81.3%), female (85.7%), never hospitalized for COVID-19 (89.3%), and received either a provider diagnosis or RT-PCR confirmed SARS-CoV-2 infection (77.1%). See Table 1 for more detail.

**Table 1.** Participant demographics. Table showing distribution of participants based on race and ethnicity, gender, age, mode of determining SARS-CoV-2 infection, and hospitalization status.

| CHARACTERISTIC | PARTICIPANTS (n=5163), No. (%) |
| --- | --- |
| **Race and Ethnicity:** |  |
| White | 4198 (81.3) |
| Hispanic or Latinx | 330 (6.4) |
| Multiracial | 159 (3.1) |
| Asian/Pacific Islander | 114 (2.2) |
| Black | 111 (2.2) |
| Hispanic or Latinx, White | 89 (1.7) |
| Black | 41 (0.8) |
| American Indian | 35 (0.7) |
| Hispanic or Latinx,<br>Multiracial | 27 (0.5) |
| Other | 22 (0.4) |
| Middle Eastern | 19 (0.4) |
| Hispanic or Latinx, American<br>Indian | 7 (0.1) |
| Hispanic or Latinx, Black | 6 (0.1) |
| Hispanic or Latinx, Other | 3 (0.1) |
| Hispanic or Latinx,<br>Asian/Pacific Islander | 2 (0.0) |
| **Gender:** |  |
| Female | 4422 (85.7) |
| Male | 714 (13.8) |
| Non-binary/non-conforming | 15 (0.3) |
| Unknown | 10 (0.2) |
| Transgender | 2 (0.0) |
| **Age:** |  |
| 18-24 | 118 (2.2) |
| 25-34 | 593 (11.5) |
| 35-44 | 1298 (25.1) |
| 45-54 | 1560 (30.2) |
| 55-64 | 1122 (21.7) |
| 65-74 | 428 (8.3) |
| 75-84 | 40 (0.8) |
| 85+ | 4 (0.1) |
| **COVID-19 Diagnosis:** |  |
| Positive COVID-19 test | 2644 (51.2) |
| Diagnosis from doctor | 1337 (25.9) |
| Self-diagnosed based on symptoms | 918 (17.8) |
| Other | 264 (5.1) |
| **Hospitalized:** |  |
| No | 4918 (83.9) |
| Yes | 876 (15.5) |
| NA | 38 (0.6) |

### Prevalence and duration of symptoms among participants with PASC

PASC individuals reported an average of 21.4 symptoms with a range of 1 to 93 symptoms occurring *(Supplemental Figure 1)*. The prevalence of symptoms is reported in Figure 1, and a graph of combined average time to symptom onset and duration among participants with PASC is reported in Figure 2. Of the 101 symptoms reported, the most prevalent symptoms included fatigue, headache, shortness of breath, difficulty concentrating, inability to exercise, cough, change in sense of taste, diarrhea, and muscle or body aches. Timing of symptom onset varied and was typically described as occurring in “waves”. Symptom duration ranged from 2 weeks to over 100 days, with changing symptoms being a prominent, long-lasting feature (Figure 2). The first wave (arrhythmia to burning calves) appeared to be heavily dominated by neurological and cardiovascular manifestations with some indicators of a strong immune response (enlarged and painful lymph nodes), followed by microvascular consequences in the second wave (Covid toes). The final wave suggested impact on endocrine (thyroid) function. At the time that they took the survey 98.0% of the respondents said they had symptoms that were unresolved, and 97.8% reported that at least one of their symptoms was intermittent, meaning it would temporarily resolve and then later return.

**Figure 1:**
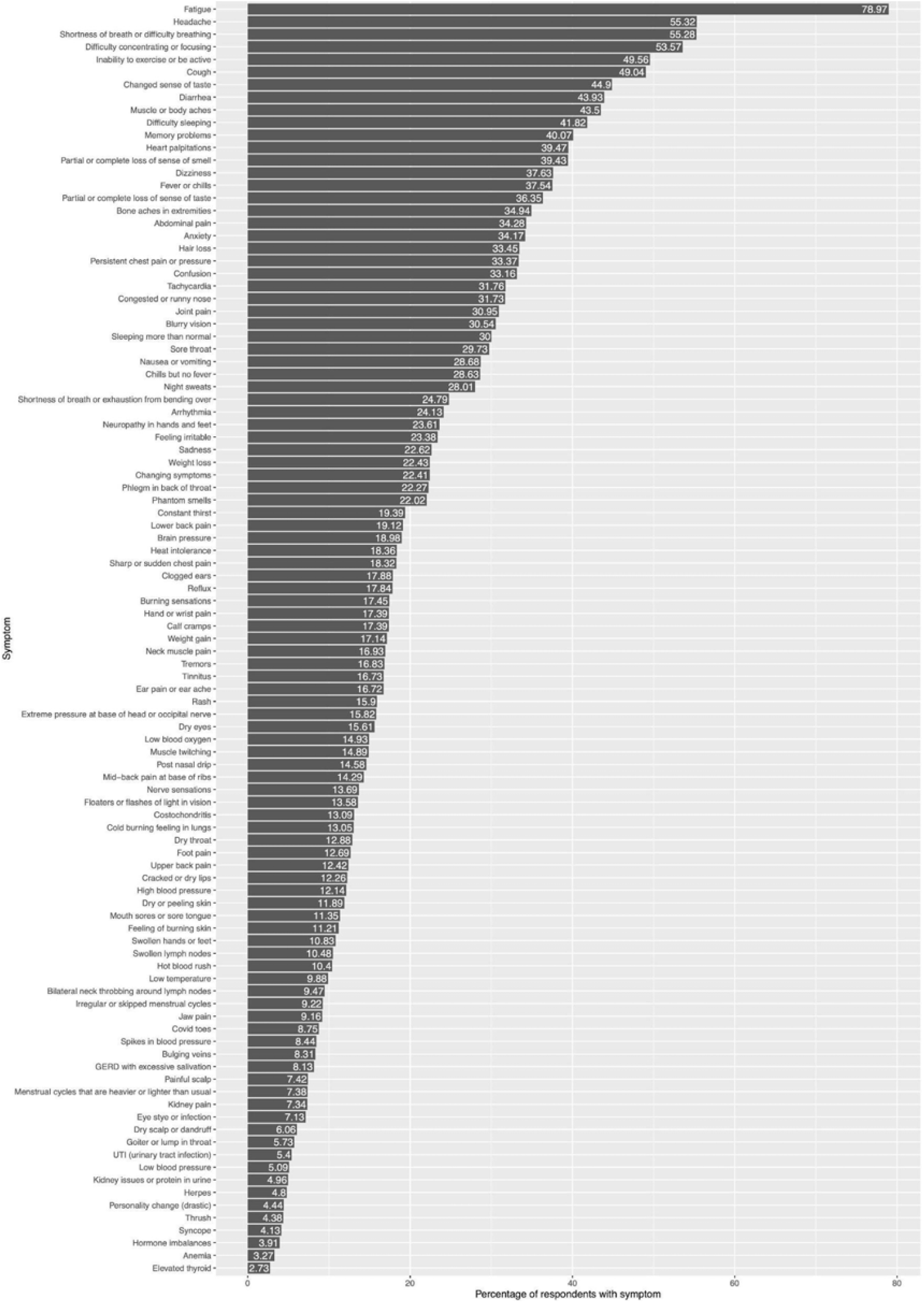
Percentage of respondents reporting each COVID-19 symptom. Graph showing percentage of participants (*x*-axis) who experienced a given symptom (*y*-axis).

**Figure 2.**
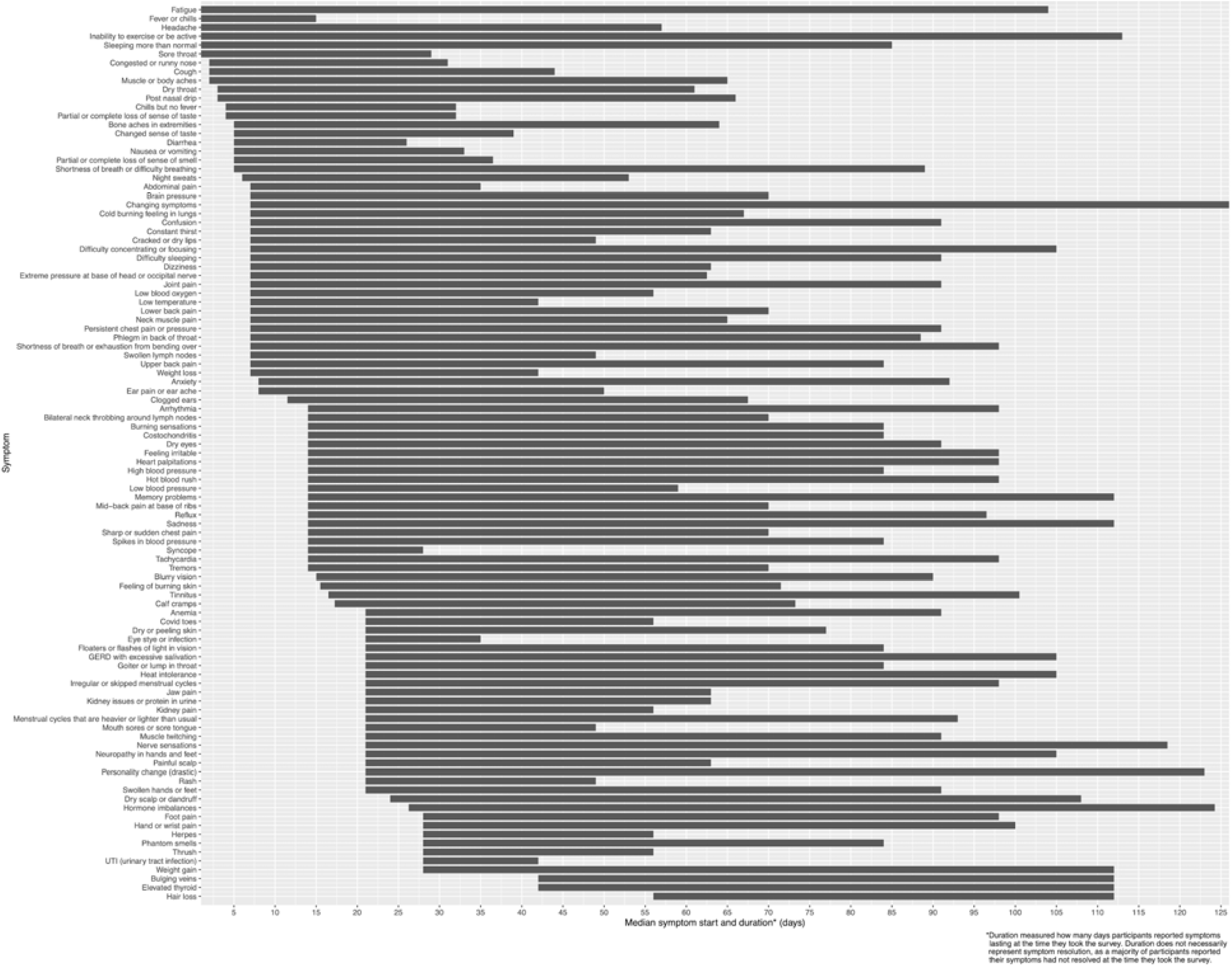
Average symptom onset and duration. Graph plotting the average time of symptom onset (in days after initial infection date) with average symptom duration. *Duration measures length of time the symptom was present and does not indicate resolution of the symptom. Participants noted these symptoms were ongoing at the time they completed the survey.

### Impact of symptoms on participants with PASC

The calculations of average time to symptom onset; symptom duration; percentage of respondents who reported the symptom as intermittent or ongoing; and the average symptom impact in terms of pain and discomfort, work impairment, and social relationship are recorded in Table 2. Variables included percentage reporting a symptom; pain and discomfort of the symptom (e.g., distress); ability to work or socialize; symptom duration; percentage reporting ongoing symptoms; and percentage reporting symptoms as intermittent. Potential morbidity, especially inability to work, was assessed through symptom severity, duration, and perceived impact on ability to work (Figure 3).

**Table 2:**
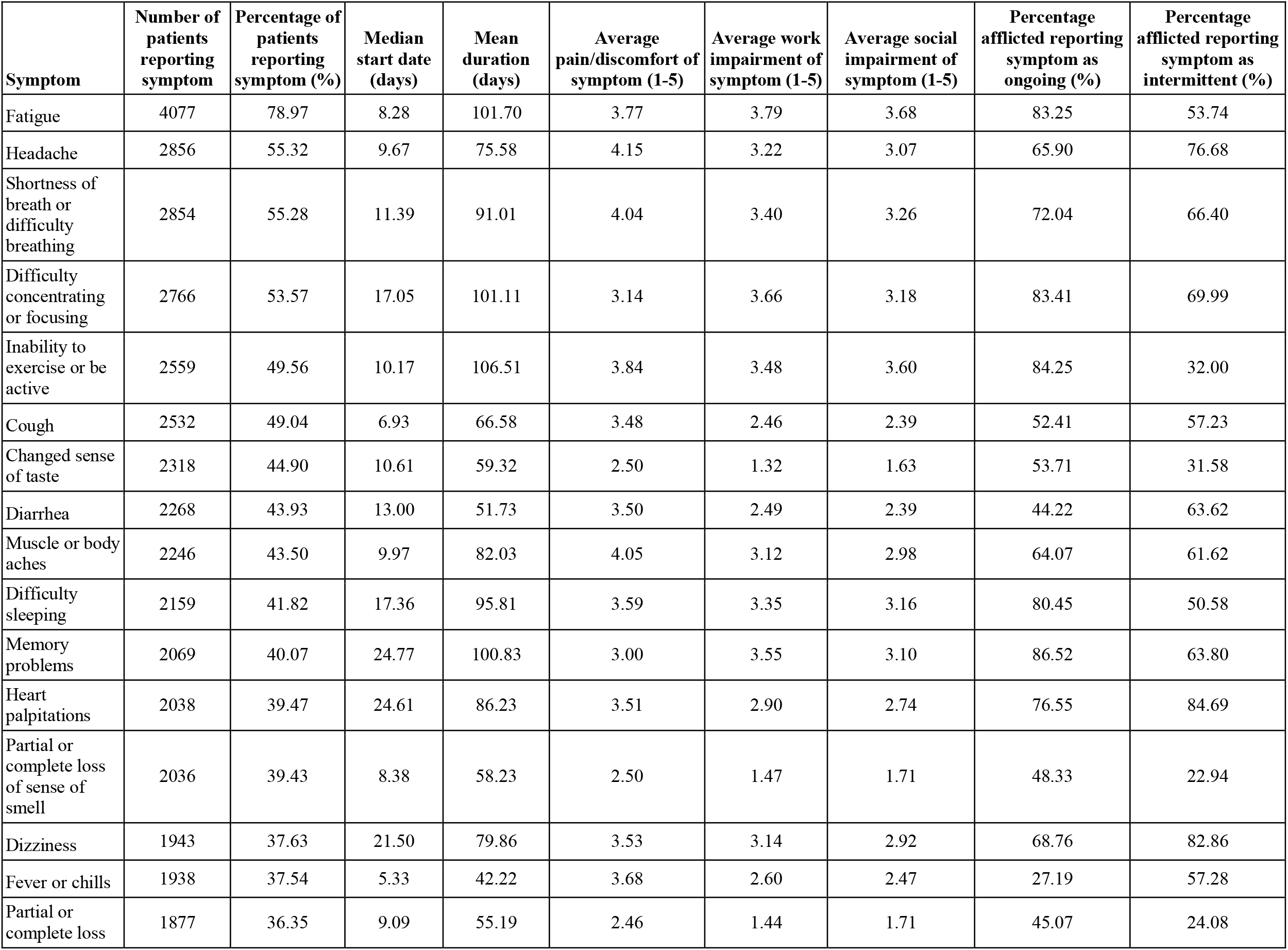
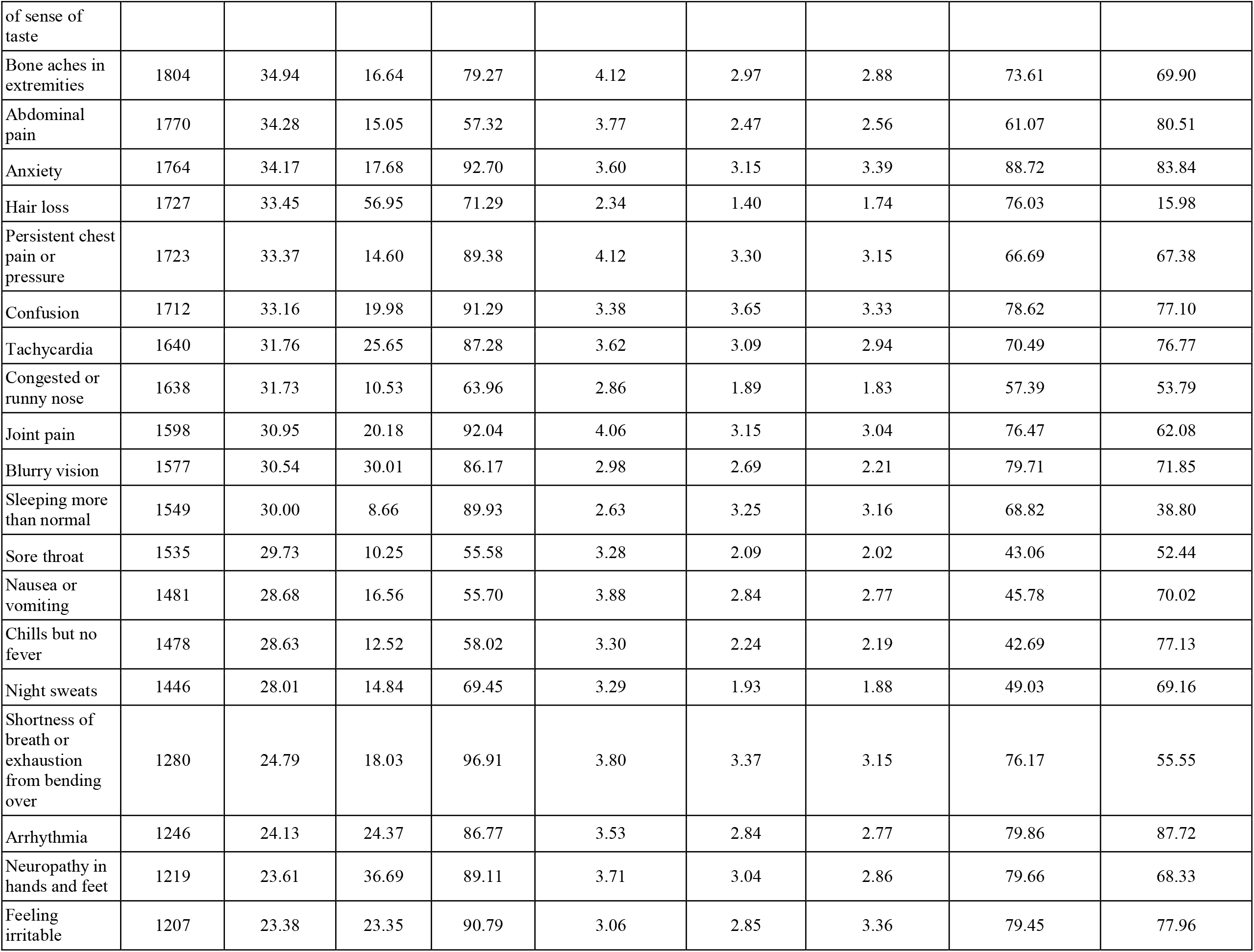
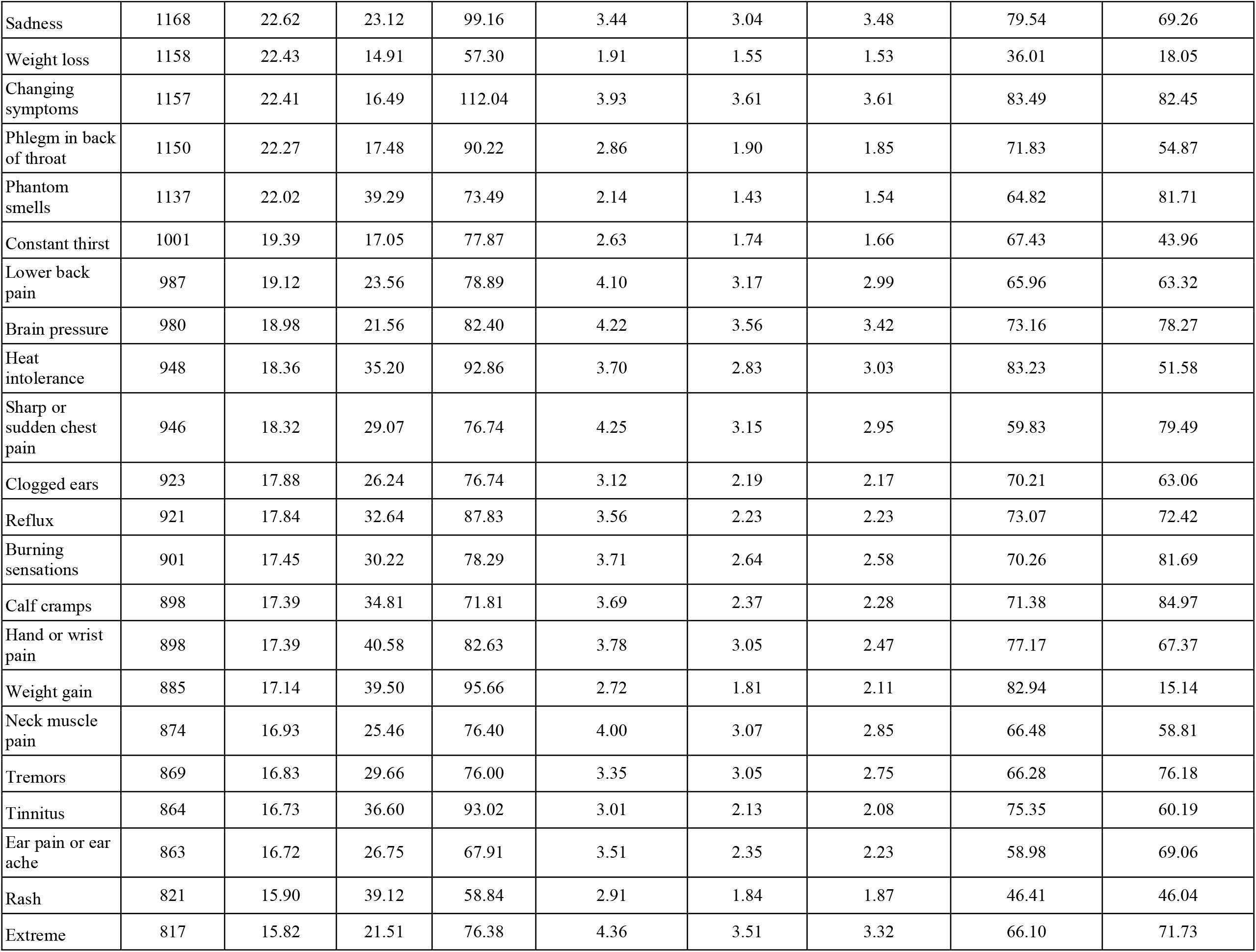
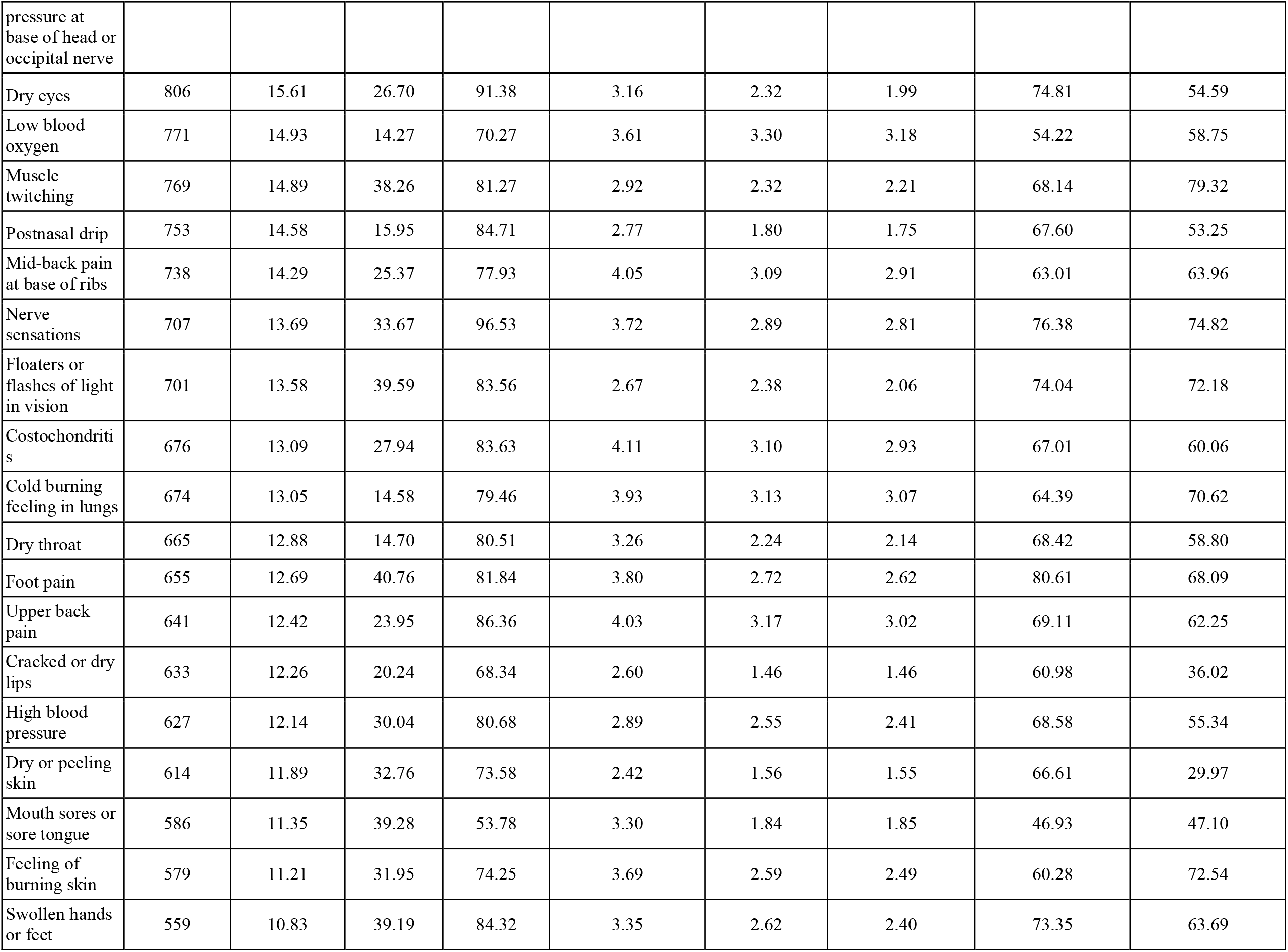
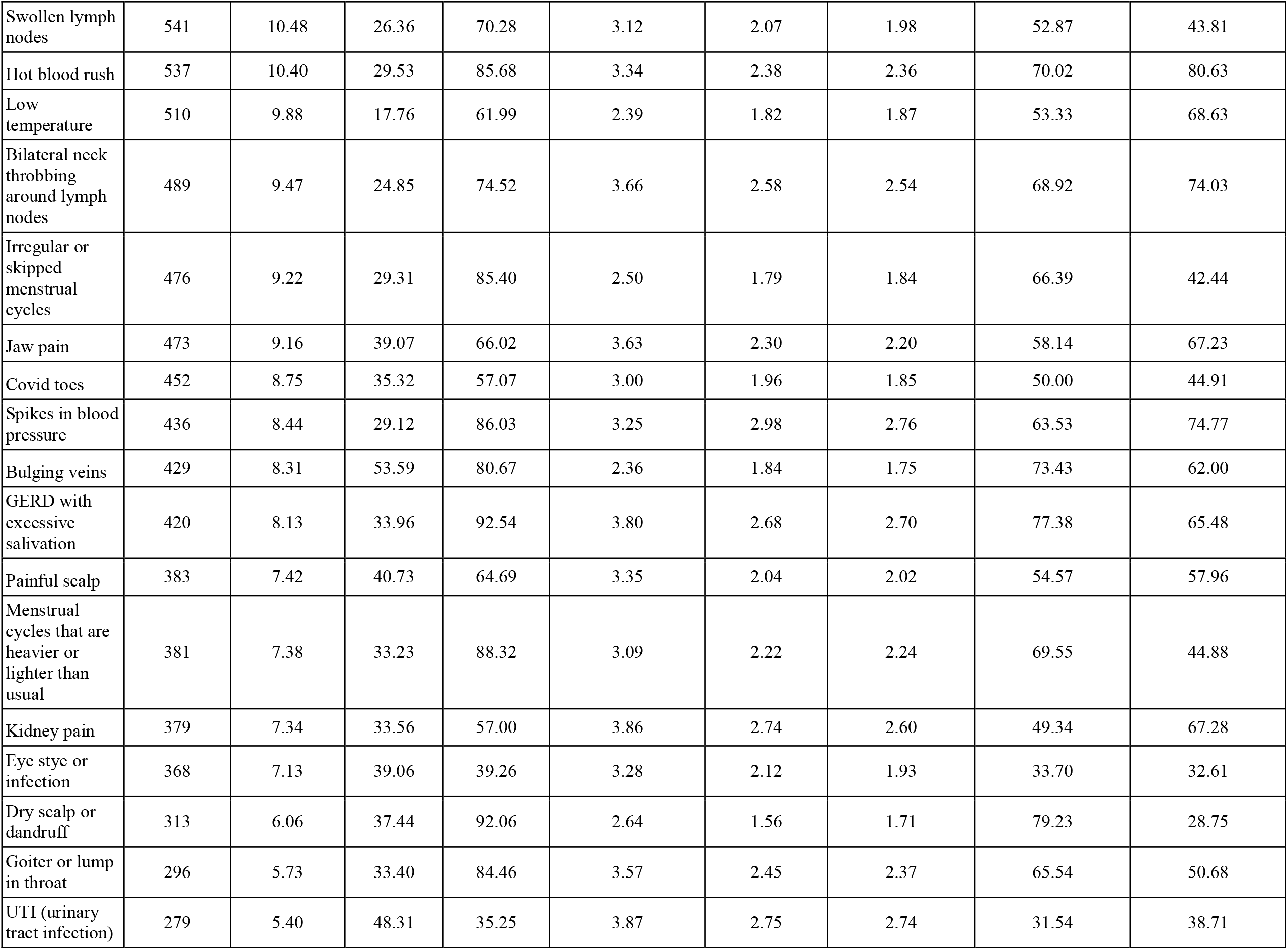
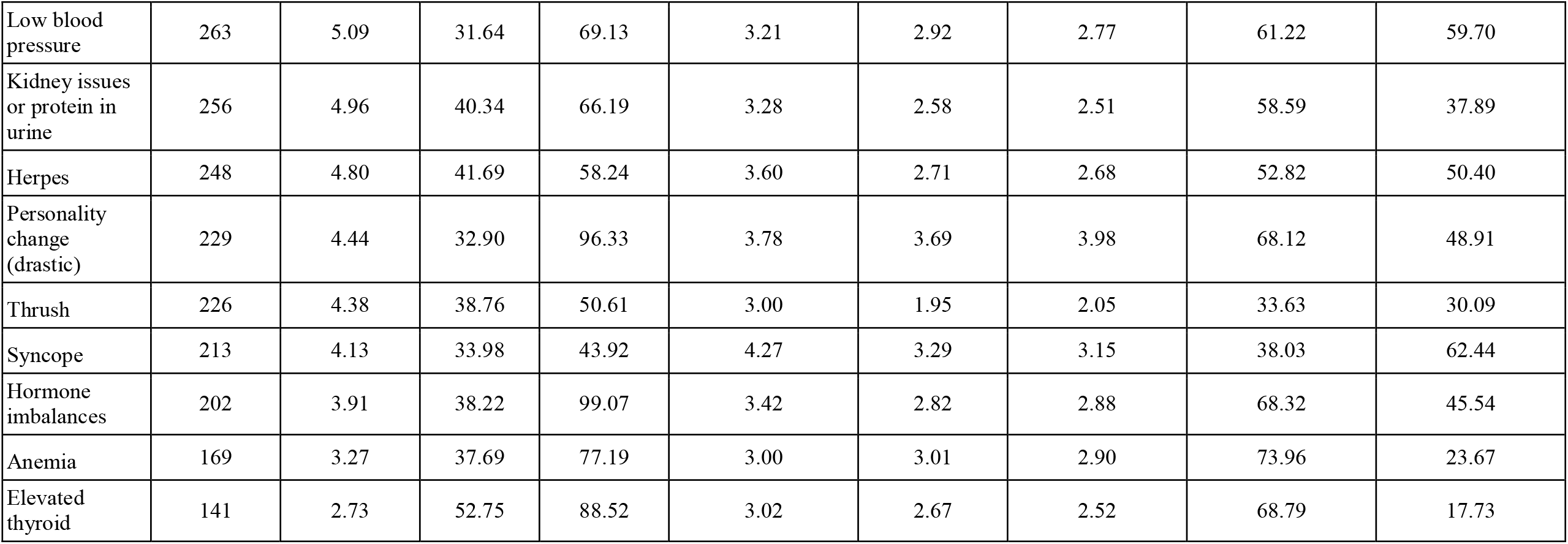
COVID-19 symptoms and their health impacts. Health impact variables include: percentage of participants reporting symptoms, pain and discomfort (e.g. level of distress), ability to work, social impairment, symptom duration, percentage reporting symptoms as ongoing, and percentage reporting symptoms as intermittent.

**Figure 3.**
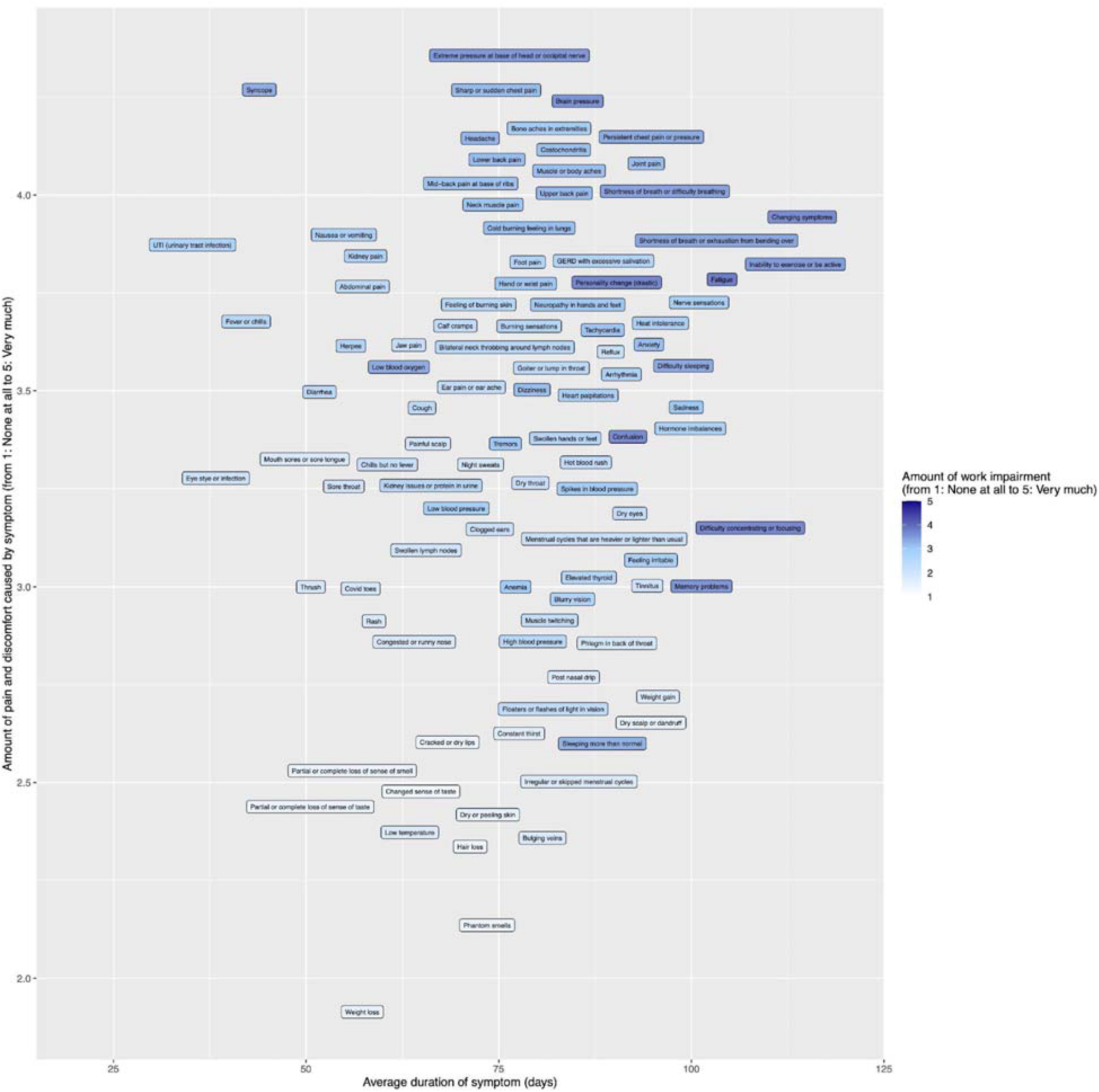
Impact of level of distress and duration of symptoms on ability to work among PASC survivors. Graph showing the effect of duration of symptoms (*x*-axis) and level of distress (*y*-axis) on ability to work. Darker shades indicate greater impact on ability to work.

Symptoms perceived to have the most impact on ability to work included fatigue, personality change, a sensation of “brain pressure”, inability to sleep, inability to exercise, difficulty concentrating, memory problems, confusion, shortness of breath, and the relapsing/remitting nature of symptoms.

## Conclusions

The current study provides much needed insight into the sequelae of acute SARS-CoV-2 infection. These findings warrant additional discussion, investigation, and context within the current knowledge of PASC. In reviewing our findings, we believe there are 4 key take-away points that warrant further discussion.

First, the fact that survey participants reported an average of 24.1 symptoms means that the average PASC patient experiences more long-lasting sequelae of COVID-19 than is currently captured by most research studies.^10, 11^ The study’s ability to detect a larger number of symptoms can be attributed to including a list of potential COVID-19 symptoms based on initial open-ended research of the range of COVID-19 sequelae conducted in July of 2020^4^ and expanded through feedback on the survey design from Survivor Corps. Strengths of this study include reporting of symptoms among the 5,163 individuals who have and are continuing to experience symptoms up to a year after SARS-CoV-2 infection, (PASC survivors). Participants were able to describe their symptoms and health impact caused by these symptoms in detail, which provides additional richness and insight into the symptom sequelae.

A second insight was that PASC may affect more women than men since 85.7% of our PASC respondents were women. Although prior research has found that older adults are at greater risk of hospitalization than younger adults, in our participant group PASC impacted people in the middle years of life most frequently with 77.0% (n= 3,980) of our participants aged 35-64. We^2^ and others^8, 12, 13^ have shown that PASC occurs more frequently in women with an age distribution similar to this study. Our participants were also largely not hospitalized, indicating a less severe initial course. 77% of participants reported clinician diagnosis or confirmed RT PCR for SARS-CoV-2, with 5% reporting “other” and ∼18% self-diagnosing. Those without confirmed diagnosis may have attributed SARS-CoV-2 infection with a confirmed antibody test that was available early in the pandemic or through the presence of COVID-specific symptoms such as loss of sense of smell. While there may be an inclination for researchers to exclude participants who do not have a positive RT PCR or antibody test from COVID-19 research, capturing the experiences of the earliest long haulers (before validated tests had become available) is essential for understanding the trajectory of PASC.

Third, participants reported multiple symptoms that appear to occur in waves. This observation is consistent with other reports, including hospitalized individuals. The temporal nature of these symptoms may provide clues into underlying pathophysiology, which is currently unclear. Others describe PASC symptoms occurring in waves which may indicate an evolving process guided by endothelial dysfunction, inflammation, oxidative stress, and hormonal imbalance.^14,15^ However, a causal relationship has not been established. These findings align with the team’s prior work in different data sets that observed medically documented symptoms in electronic health records longitudinally and established a temporal order and inclusion of only new symptoms at 0-11 and 60+days following a positive SARS-CoV-2 PCR test. In this work we identified 5 symptom clusters in non-hospitalized PASC survivors, with dominant symptoms in those clusters all represented in the top quartile of symptoms reported here. Of note is the changing nature of symptoms, which appears to be a particularly distressing feature among PASC survivors.

Finally, PASC appears to exert variable degrees of impact on individuals, but mechanistic insight is lacking. Most studies to date have focused on hospitalized patients; however, in our study most participants were never hospitalized, suggesting that PASC has profound effects, independent of COVID-19 severity. Factors that were reported to have the most impact on participants’ ability to work after developing PASC included the relapsing/remitting nature of symptoms, the long duration of many symptoms reported as unresolved, as well as fatigue and symptoms associated with altered cognition or memory impairment. Overall, reported symptoms varied with regards to duration and for their degree of distress, suggesting that a symptom of PASC can cause severe health impacts for some and milder impacts for others, making PASC an experience that is unique to the individual. Considering the great number and severity of health impacts caused by PASC, future research with PASC patients should endeavor to collect qualitative data, asking patients to report on the lasting impacts including their ability to work, maintain social relationships, their mental health, as well as basic everyday functioning. Analysis and fusion of biological and qualitative data are essential to ensure that health policy can be designed to meet PASC survivors’ needs.

## Limitations

The sequelae of acute SARS-CoV-2 infection are just beginning to be understood, and it is clear that PASC affects those who were never hospitalized for severe illness. More work is needed to corroborate our findings and to document the temporal nature of symptoms of PASC. Further research is needed to determine non-modifiable risk factors associated with increased risk of acquiring PASC, as well as aberrant innate and adaptive immune responses associated with PASC.

## Data Availability

The data that support the findings of this study are available from the corresponding author, NL, upon reasonable request.

## Acknowledgements

Thank you to Survivor Corps for mobilizing many thousands of COVID-19 survivors to participate in research to find a cure for COVID-19, and for being the epicenter of hope for so many. We would like to express our deep gratitude to the thousands of long haulers who spent many hours taking this survey while suffering from the impacts of PASC. The answers we need to cure PASC will come from listening and learning from those who suffer from the disease.

We would also like to thank the Precision Health Initiative at Indiana University for their support on this project.

## Table and Figure Legends

**Supplemental Figure 1.**
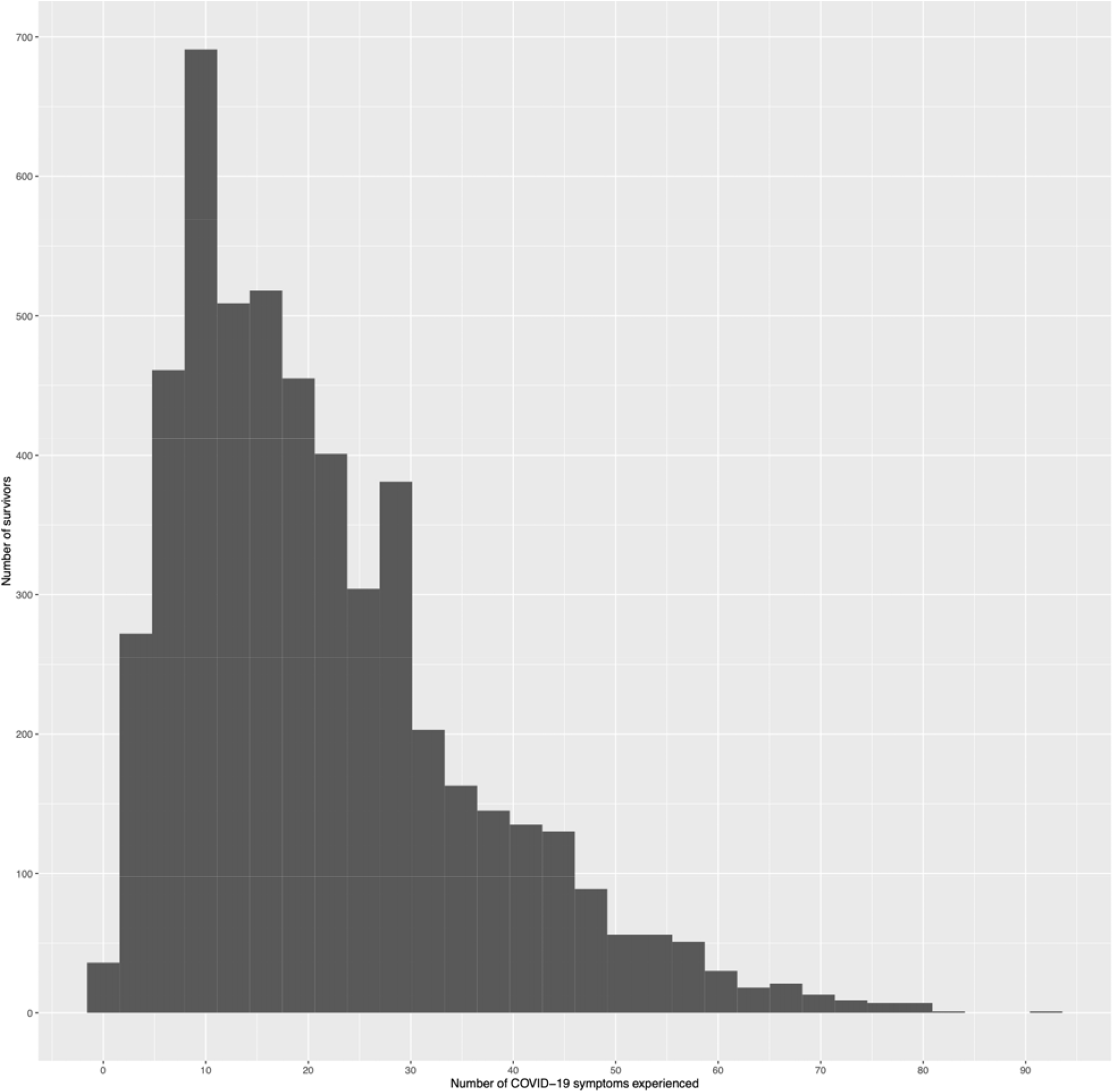
Distribution of the number of symptoms reported by participants. Graph visualizing the distribution of the number of symptoms reported by survey participants.

## References

1. Nehme M, Braillard O, Alcoba G, et al. COVID-19 Symptoms: Longitudinal Evolution and Persistence in Outpatient Settings. Ann Intern Med. [Epub ahead of print 8 December 2020].DOI:10.7326/M20-5926

2. Huang, Y, Pinto, MD, Borelli, JL, et al. COVID Symptoms, Symptom Clusters, and Predictors for Becoming a Long-Hauler: Looking for Clarity in the Haze of the Pandemic. medRxiv 2021.03.03.21252086; doi: https://doi.org/10.1101/2021.03.03.21252086

3. Johns Hopkins University of Medicine. COVID-19 Dashboard by the Center for Systems Science and Engineering (CSSE) at Johns Hopkins University (JHU). Johns Hopkins University of Medicine Coronavirus Resource Center. January 22, 2020. accessed March 9, 2021. https://coronavirus.jhu.edu/ma p.html

4. Lambert, N. J. & Survivor Corps COVID-19 “Long Hauler” Symptoms Survey Report. Indiana University School of Medicine. Published online July 7, 2020. accessed March 19, 2021. https://static1.squarespace.com/static/5e8b5f63562c031c16e36a93/t/5f459ef7798e8b6037fa6c57/1598398215120/2020+Survivor+Corps+COVID-19+%27Long+Hauler%27+Symptoms+Survey+Report+%28revised+July+25.4%29.pdf

5. Logue JK, Franko NM, McCulloch DJ, et al. Sequelae in Adults at 6 Months After COVID-19 Infection. JAMA Netw Open. 2021;4(2):e210830. DOI:10.1001/jamanetworkopen.2021.0830

6. Garrigues E, Janvier P, Kherabi Y, et al. Post-discharge persistent symptoms and health-related quality of life after hospitalization for COVID-19. J Infect. 2020;81(6):e4–e6. DOI:10.1016/j.jinf.2020.08.029

7. Tenforde M, Kim S, Lindsell C, et al. 6-month consequences of COVID-19 in patients discharged from hospital: a cohort study. Lancet. 2021;397(10270): 220–232. DOI:10.1016/S0140-6736(20)32656-8

8. Carfi, A. Bernabei, R., Landi, F., et al. Persistent symptoms in patients after acute COVID-19. JAMA. 2020;324(6): 603–605. DOI:10.1001/jama.2020.12603

9. Alwan NA. Surveillance is underestimating the burden of the COVID-19 pandemic. Lancet. 2020;396(10252): e24. DOI:10.1016/S0140-6736(20)31823-7

10. Tenforde MW, Kim SS, Lindsell CJ, et al. Symptom Duration and Risk Factors for Delayed Return to Usual Health Among Outpatients with COVID-19 in a Multistate Health Care Systems Network — United States, March–June 2020. MMWR Morb Mortal Wkly Rep. 2020;69:993–998. DOI: http://dx.doi.org/10.15585/mmwr.mm6930e1

11. Stavem K, Ghanima W, Olsen MK, et al Persistent symptoms 1.5–6 months after COVID-19 in non-hospitalised subjects: a population-based cohort study. Thorax. 2021;76:405–407.

12. Sudre, C.H., Murray, B., Varsavsky, T. et al. Attributes and predictors of long COVID. Nat Med. 2021. https://doi.org/10.1038/s41591-021-01292-y

13. Davido B, Seang S, Tubiana R, Truchis P. Post–COVID-19 chronic symptoms: a postinfectious entity?. Clin Microbiol Infect. 2020; 26(11): 1448–1449. https://doi.org/10.1016/j.cmi.2020.07.028

14. Theoharides TC, Conti P. COVID-19 and multisystem inflammatory syndrome, or is it mast cell activation syndrome?. J Biol Regul Homeost Agents. 2020 Sep 1;34(5).

15. Leung TYM, Chan AYL, Chan EW, et al. Short-and potential long-term adverse health outcomes of COVID-19: a rapid review. Emerg Microbes Infect. 2020; 9:1, 2190–2199, DOI: 10.1080/22221751.2020.1825914

